# Evidence that minocycline treatment confounds the interpretation of neurofilament as a biomarker

**DOI:** 10.1101/2024.05.01.24306384

**Authors:** Juliana E Gentile, Christina Heiss, Taylor L Corridon, Meredith A Mortberg, Stefanie Fruhwürth, Kenia Guzman, Lana Grötschel, Kwan Chan, Neil C Herring, Timothy Janicki, Rajaa Nhass, Janani Manavala Sarathy, Brian Erickson, Ryan Kunz, Alison Erickson, Craig Braun, Katherine T Henry, Lynn Bry, Steven E Arnold, Eric Vallabh Minikel, Henrik Zetterberg, Sonia M Vallabh

## Abstract

Neurofilament light (NfL) concentration in cerebrospinal fluid (CSF) and blood serves as an important biomarker in neurology drug development. Changes in NfL are generally assumed to reflect changes in neuronal damage, while little is known about the clearance of NfL from biofluids. We observed an NfL increase of 3.5-fold in plasma and 5.7-fold in CSF in an asymptomatic individual at risk for genetic prion disease following 6 weeks’ treatment with oral minocycline for a dermatologic indication. Other biomarkers remained normal, and proteomic analysis of CSF revealed that the spike was exquisitely specific to neurofilaments. NfL dropped nearly to normal levels 5 weeks after minocycline cessation, and the individual remained free of disease 2 years later. Plasma NfL in dermatology patients was not elevated above normal controls. Dramatically high plasma NfL (>500 pg/mL) was variably observed in some hospitalized individuals receiving minocycline. In mice, treatment with minocycline resulted in variable increases of 1.3- to 4.0-fold in plasma NfL, with complete washout 2 weeks after cessation. In neuron-microglia co-cultures, minocycline increased NfL concentration in conditioned media by 3.0-fold without any visually obvious impact on neuronal health. We hypothesize that minocycline does not cause or exacerbate neuronal damage, but instead impacts the clearance of NfL from biofluids, a potential confounder for interpretation of this biomarker.

## Introduction

Neurofilament proteins light, medium, and heavy (NfL, NfM, and NfH) are components of an intermediate filament that structurally supports neurons^1^. As neuronal damage causes leakage of these proteins from the cytosol into biofluids, NfL in cerebrospinal fluid and blood are elevated, to various degrees, in virtually all neurological disorders^2, 3^. Although NfL is not diagnostically specific, it can potentially provide information on patient prognoses and treatment responses^4^. As a result, in the development of tofersen for *SOD1* ALS, NfL has been used both as an Accelerated Approval endpoint in symptomatic patients^5^ and as a criterion for randomization of pre-symptomatic patients to drug or placebo^6^. Conversely, increased NfL has been cited as one reason for premature termination of the phase II trial of branaplam in Huntington’s disease^7, 8^. In principle, the concentration of any biomarker analyte is a function of both the rate of its production in tissue and release into fluids, as well as its catabolism and/or clearance from fluids. Yet the above examples illustrate that NfL is widely used and interpreted as a marker of neurological insult, while little is known about how it is removed from biofluids.

Minocycline is a tetracycline antibiotic used clinically both for infections and for dermatologic conditions^9–11^. It has been shown in rodents and in humans to suppress microglial activation and proliferation^12–14^. This finding has led to clinical trials of minocycline in diverse neurologic indications, though the results have generally been null or unfavorable^15, 16^. In clinical trials of traumatic brain injury (TBI) and multiple sclerosis clinically isolated syndrome (MS-CIS), randomization to minocycline was associated with a ∼3-fold increase in plasma NfL, which was interpreted as meaning that the drug may have contributed to neurodegeneration or was intrinsically neurotoxic^14, 17^.

Prion disease exhibits a more dramatic elevation of NfL than that seen in perhaps any other neurological disorder^18–20^. We and others have followed pre-symptomatic people at risk for genetic prion disease, however, and have found that plasma NfL increases in only a brief window prior to the onset of symptom^20–23^, consistent with the rapid clinical course of prion disease^24^. Here, we report a healthy, at-risk research participant with a transient 3.5-fold spike in plasma NfL and a 5.7-fold spike in CSF NfL following a 6-week course of minocycline, in the absence of any other biomarker signs of prion disease and without subsequent onset of symptoms. Supportive but inconsistent results from studies of discarded clinical plasma samples, administration of minocycline to mice, and minocycline treatment of neuron-microglia co-cultures lead us to hypothesize that minocycline affects NfL by inhibiting its clearance, posing a potential confounder for the interpretation of this important biomarker.

## Results

An asymptomatic cohort study participant under age 50 harboring a *PRNP* mutation exhibited a spike in NfL concentration in both plasma and CSF unaccompanied by any change in other fluid biomarkers of prion disease (Table 1). At this same study visit, this individual reported having just completed, 4 days earlier, a 6- week course of 200 mg/day oral minocycline for a dermatologic indication. NfL levels dropped substantially 5 weeks after minocycline cessation and were normal 1 year later. This individual has not developed symptoms of prion disease in >2 years of follow-up after the NfL spike. Based on plasma NfL increases in trials of minocycline in TBI and MS-CIS^14, 17^, we speculated that minocycline might have been the cause of the biofluid NfL increase in our study participant.

**Table 1.**
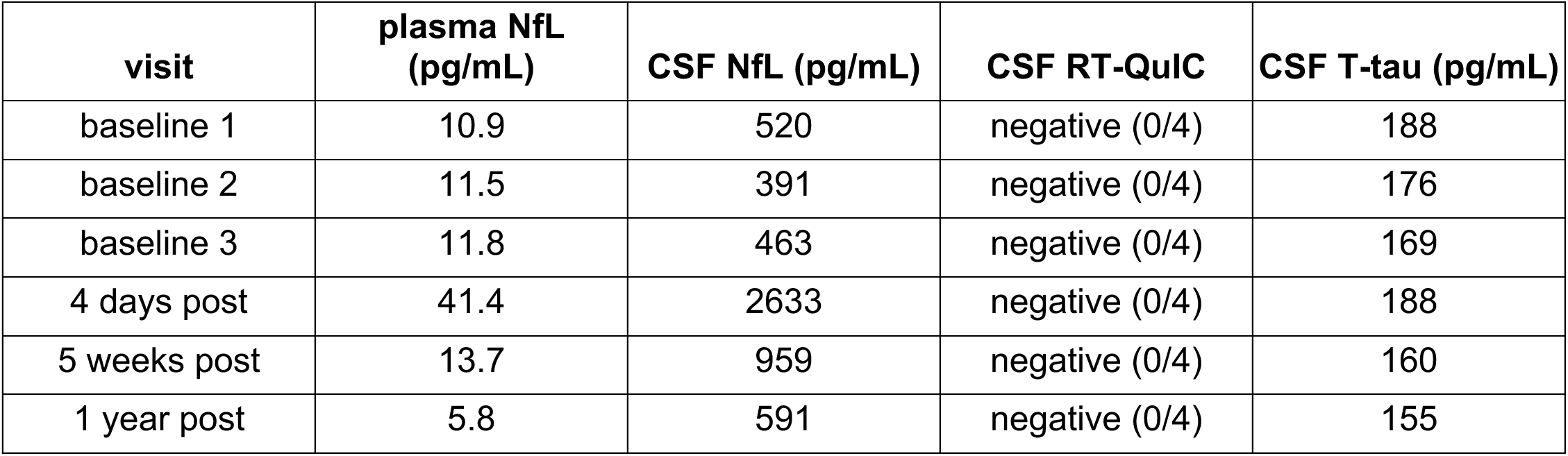
Fluid biomarker values in a cohort study participant before and after taking minocycline. Baselines were taken over a >3 year period prior to minocycline treatment. “Post” indicates time post cessation of minocycline. Parenthetical RT-QuIC values indicate positive replicates over total replicates.

We performed proteomics using tandem mass tags (TMT) on duplicate CSF samples from this individual’s first 5 serial lumbar punctures (Figure 1). Of 1,190 proteins quantified, increases at 4 days post-minocycline were exquisitely specific to NfL (*NEFL,* 1 unique peptide) and NfM (*NEFM,* 5 unique peptides) (Figure 1A, Table S1, S2), with smaller changes in microglial proteins TREM2 and AIF1, and a reduction in intermediate filament VIM. By TMT, all of these proteins returned to their baseline levels within 5 weeks (Figure 1B).

**Figure 1.**
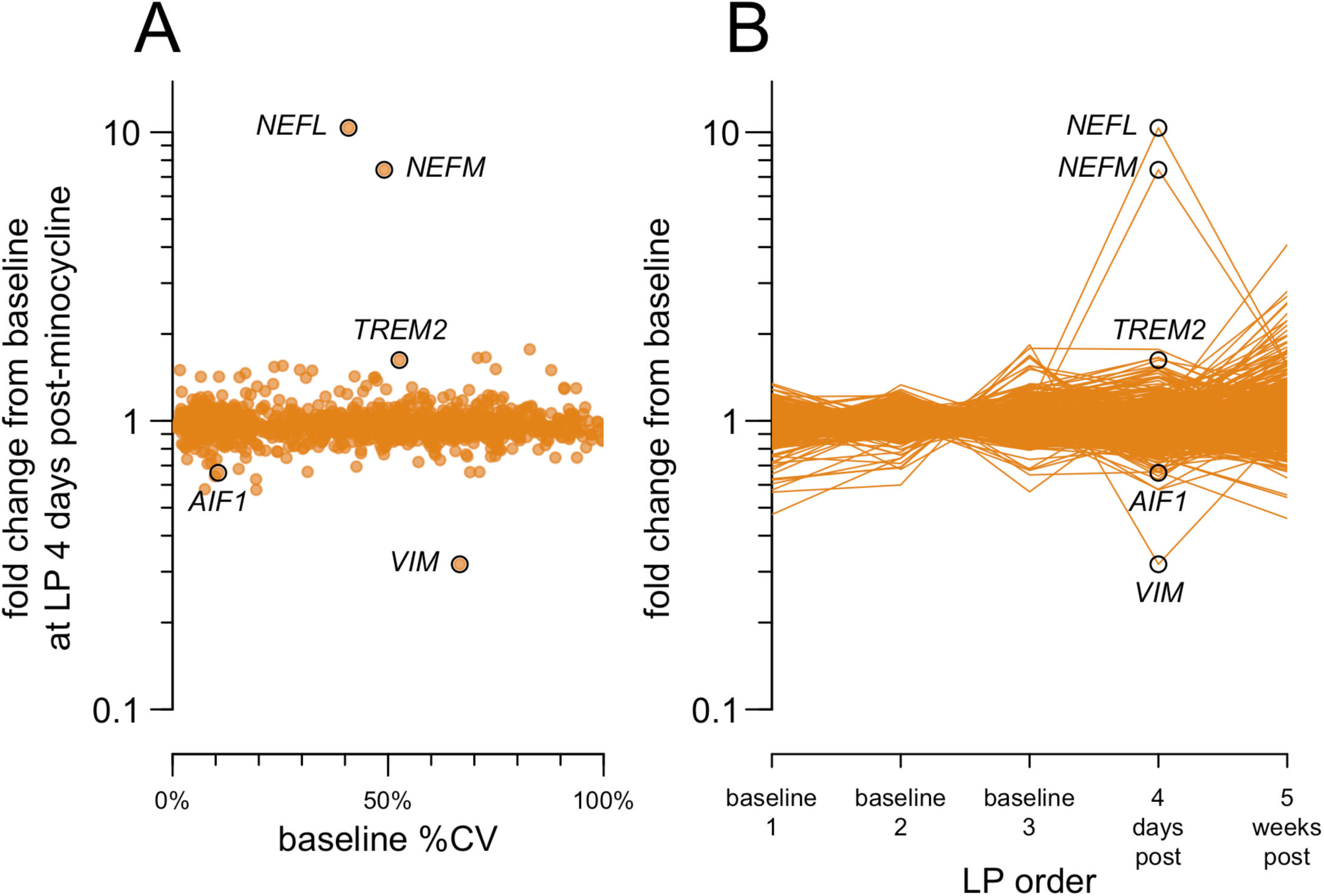
Proteomic analysis of serial CSF samples from the minocycline-treated index case. CSF samples were analyzed by TMT in technical duplicate in a single plex. Of 6,705 peptides quantified (Table S1), 6,588 with technical replicate coefficient of variation (CV) <25% were grouped by gene symbol to yield relative protein abundance (Table S2). Of 1,215 gene symbols encoding detected peptides, 1,190 had a coefficient of variation among the three baseline samples (baseline %CV) of <100% and were analyzed here. **A)** Coefficient of variation among the three baseline samples (baseline %CV) versus fold change at 4 days post-minocycline relative to the baseline mean. **B)** Ratio of each sample’s abundance to the mean of the three baseline visits.

Although our index case harbors a pathogenic *PRNP* mutation, the lack of manifest disease 2 years after the NfL spike led us to wonder whether minocycline would elicit an increase in biofluid NfL levels in individuals lacking any predisposition to neurodegeneration. We therefore collected discarded plasma samples from dermatology patients with diagnoses of acne, rosacea, eczema, or dermatitis — reasoning that such patients are generally otherwise healthy — who had a new prescription start in the past 30 days (Figure 2A, Table S3-S4) and had an order for complete blood count (CBC) to allow collection of the excess whole blood after completion of clinical testing. We also considered well visit controls unselected for starting any medications. However, as blood testing is not routinely ordered in outpatient dermatology visits, our sample collection was inherently limited to individuals who happened to have had a blood draw for any other reason around the time of their dermatology visit.

**Figure 2.**
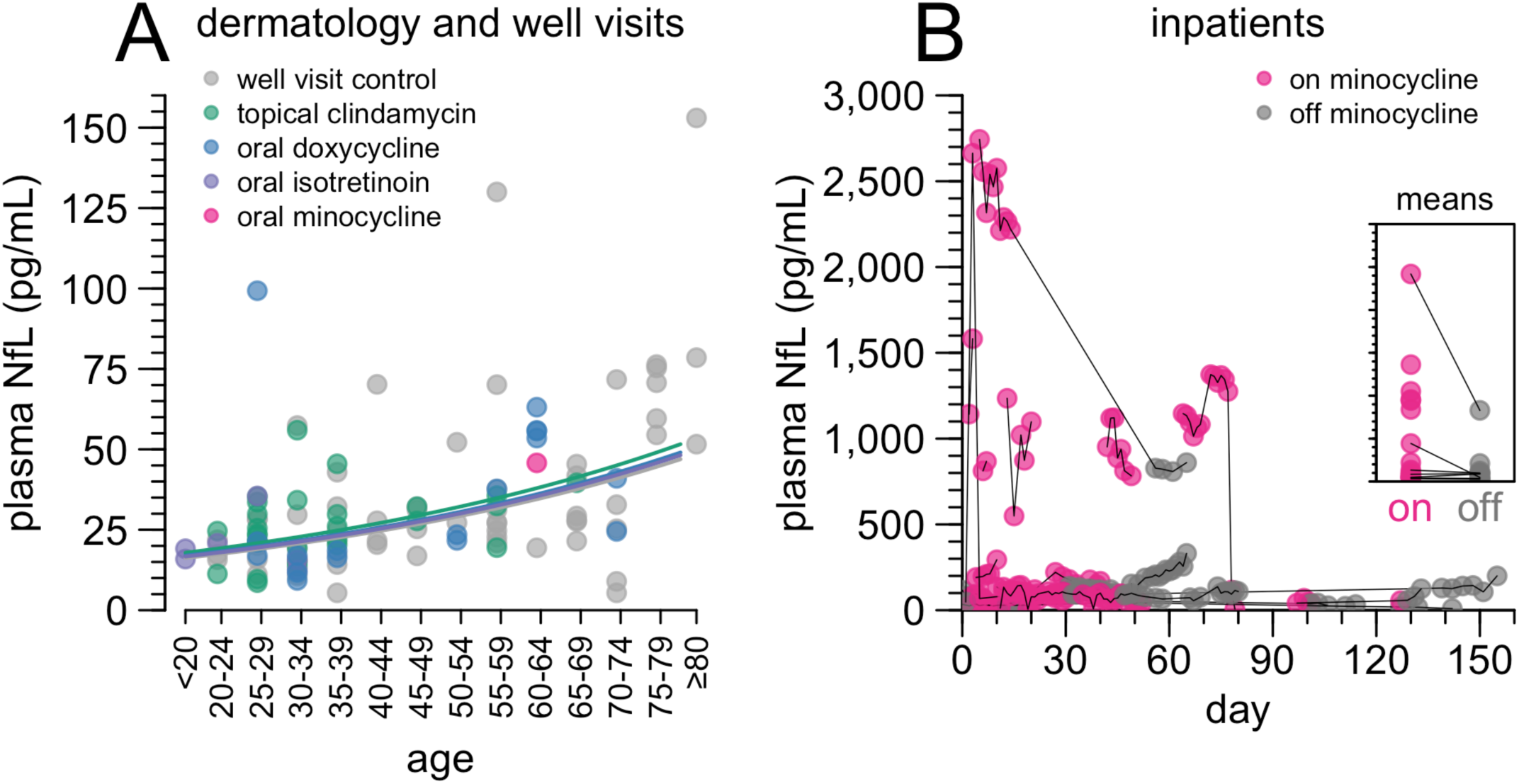
Plasma NfL levels in discarded plasma cohorts. **A)** Plasma NfL in control patients with a recent well visit (gray) or with a recent dermatology encounter, diagnosis of acne, rosacea, dermatitis, or eczema, and a recent prescription start for topical clindamycin (green), oral doxycycline (blue), oral isotretinoin (purple), or oral minocycline (magenta). For detailed inclusion criteria, see Methods. Points represent individual patients; where multiple blood samples were available for one patient, the mean plasma NfL value is plotted. Curves represent log-linear regression best fits for each group with N > 1. For individual values and model fits see Table S4-S5. **B)** Plasma NfL in inpatients prescribed minocycline. Points represent individual blood samples, thin black lines connect serial samples from the same patient. Magenta points are timepoints when on drug and following at least 7 continuous days on drug, all other points are gray. The inset panel shows mean on and mean off values for each patient, with thin lines connecting on/off means for the same patient. Individual NfL trajectories and times on/off drug for each inpatient are shown in Figure S1. Inpatient details are provided in Table S6-S8.

Samples from the entire discarded plasma cohort generally exhibited higher NfL values than seen in our own prion disease cohort study or in established reference ranges^25^ (Figure 2A). This finding was observed across all groups including well visit controls, and thus may be pre-analytical in nature, though NfL is reported to be robust to pre-analytical variables^3, 26, 27^ and we were unable to identify anything anomalous in the sample handling protocol (Methods). The higher-than-typical readings corresponded to higher baselines across all age groups, with a mean plasma NfL level of 16.7 pg/mL at 18 years of age. The relationship between NfL and age (+1.7% per year of life, log-linear regression, Table S5) was not higher than in previous reports^25, 26, 28^. Unfortunately, over an 18 month window for prospective sample collection, we obtained plasma from only 1 dermatology patient with a recent oral minocycline start; this one individual’s plasma NfL was not anomalous relative to other samples in this collection. NfL levels in dermatology patients receiving oral doxycycline or isotretinoin, or topical clindamycin, were not significantly different from well visit controls, arguing that these dermatological conditions do not themselves increase NfL (Figure 2A).

We also collected discarded plasma samples from hospital inpatients who received minocycline. As these patients had disparate diagnoses, reasons for hospitalization (Table S6) and reasons for minocycline administration, we used these individuals as their own controls, comparing plasma NfL in blood draws collected while on and off minocycline (Figure 2B). Samples from patients receiving minocycline were not significantly higher than during periods of time during which they were not receiving the drug (P = 0.059, partially paired Wilcoxon test). A subset of patients demonstrated extremely high (>500 pg/mL) NfL while receiving minocycline, a level not observed in any off-minocycline samples, except for one individual who dropped from a mean of 2,418 pg/mL while receiving minocycline, to 830 pg/mL after completing minocycline therapy. Within our inpatient cohort, no specific clinical co-variates were identified with individuals with ultra-high NfL while receiving minocycline — for instance, not all had neurological conditions (Table S6-S8).

To fill gaps in the clinical cohorts to power specific questions of NfL levels relative to minocycline exposure, we established a rodent model to determine minocycline’s effects on plasma NfL. To establish a baseline, we serially sampled plasma and measured NfL in C57BL/6N mice aged 6-22 weeks (N=272 samples total; Figure 3A, Table S9). Plasma NfL is both higher and more variable in male than in female mice (mean 167 vs. 86 ng/ml, mean CV 97% vs. 46% across all samples; Figures 3B-C, Table S10). In males, inter-animal variability was higher than within-animal variability over time (CV = 62% vs. 48%, Figure 3D-E) but this was not the case in females (CV = 23% between-animal and 37% within-animal, Figure 3D-E). This finding suggested that mouse studies with plasma NfL as the primary outcome are better powered with female mice than male or mixed-sex cohorts. In addition, study designs comparing animals to their own baseline do not necessarily have any advantage over study designs comparing animals to one another.

**Figure 3.**
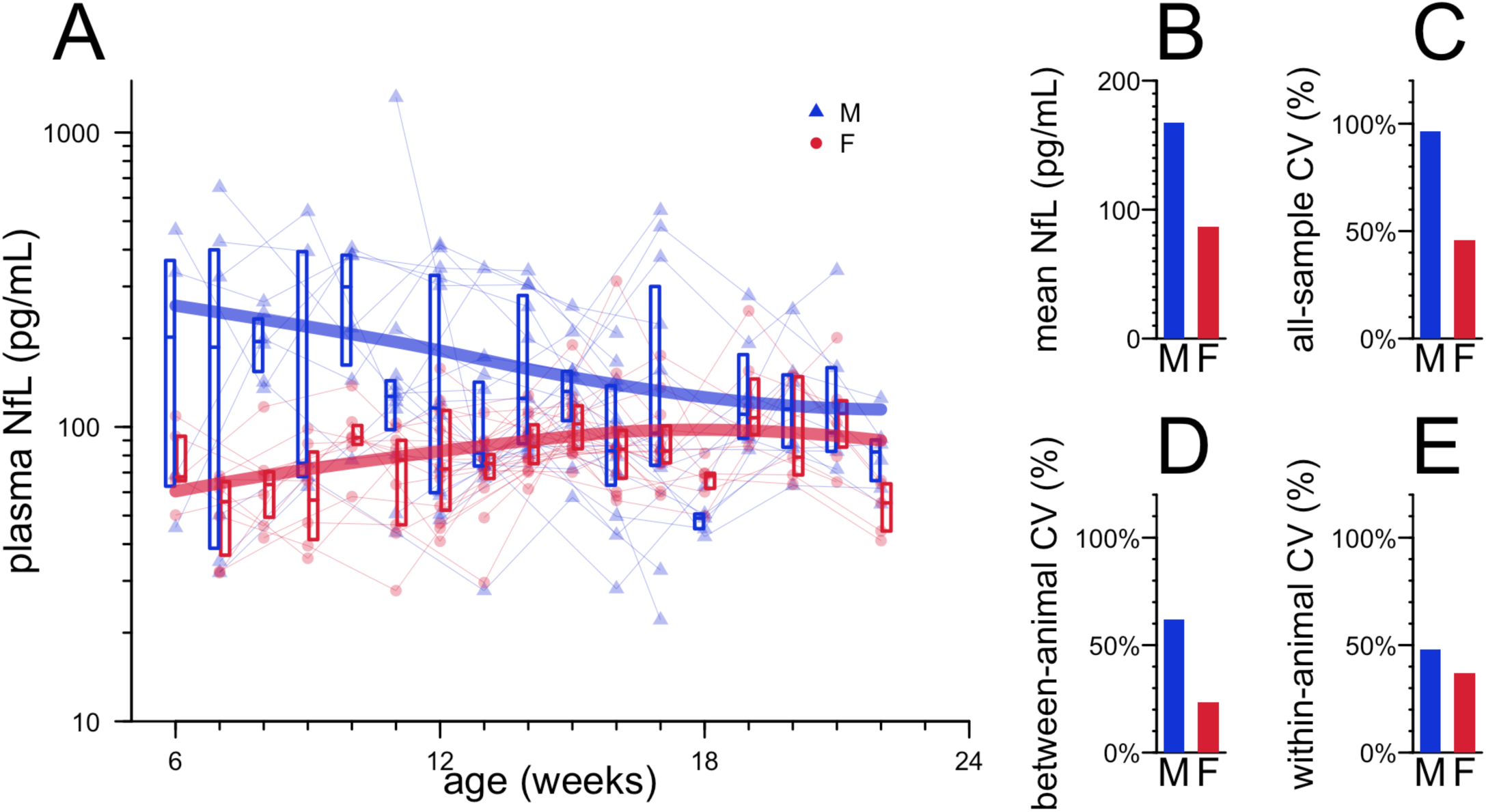
Natural history of plasma NfL in naive C57BL/6N mice. **A)** Plasma NfL values from N=272 serial bleeds. Points represent individual animals (blue triangles for males, red circles for females), thin lines connect serial samples from individual animals, boxes represent medians and interquartile ranges by sex and timepoint, and thick bars represent locally estimated scatterplot smoothing (LOESS) fits by sex. **B)** Mean NfL values by sex across all animals, all timepoints. **C)** Coefficient of variation by sex across all samples, all timepoints. **D)** Mean coefficient of variation by sex, between animals within each timepoint. **E)** Mean coefficient of variation by sex, within animal between timepoints.

Minocycline has a shorter half-life in rodents than in humans, such that a 50 mg/kg dose in mice is estimated to provide exposure similar to the common clinically used dose of minocycline, 200 mg daily, which corresponds to ∼3 mg/kg for a 70 kg human^29–31^. We treated N=16 mixed-sex mice per arm with 50 mg/kg minocycline intraperitoneally (i.p.) once daily. After 6 days of dosing, plasma NfL was 4.0 times higher in minocycline-treated mice than untreated mice (median 1211 vs. 304 pg/mL, P = 1.8e-6, Wilcoxon test, Figure 4A, Table S12-S14). This difference grew further by 13 days (median 1169 vs. 214 pg/mL, P = 3.6e-6, Figure 4A). However, this dose was not well-tolerated over this length of time: by day 10, the average minocycline-treated mouse had lost 6% body weight, and some began to require euthanasia (Figure S2A, Table S15-S16). Dosing was therefore discontinued after day 15; plasma NfL in the formerly minocycline-treated animals then dropped rapidly and was not significantly higher than the untreated animals thereafter (Figure 4A). Bulk RNA-seq on the cortex of minocycline-treated versus untreated animals identified several downregulated microglial genes but also a large number of apparently non-specific hits that may reflect cellular distress or death (Figure S2B, Table S17).

**Figure 4.**
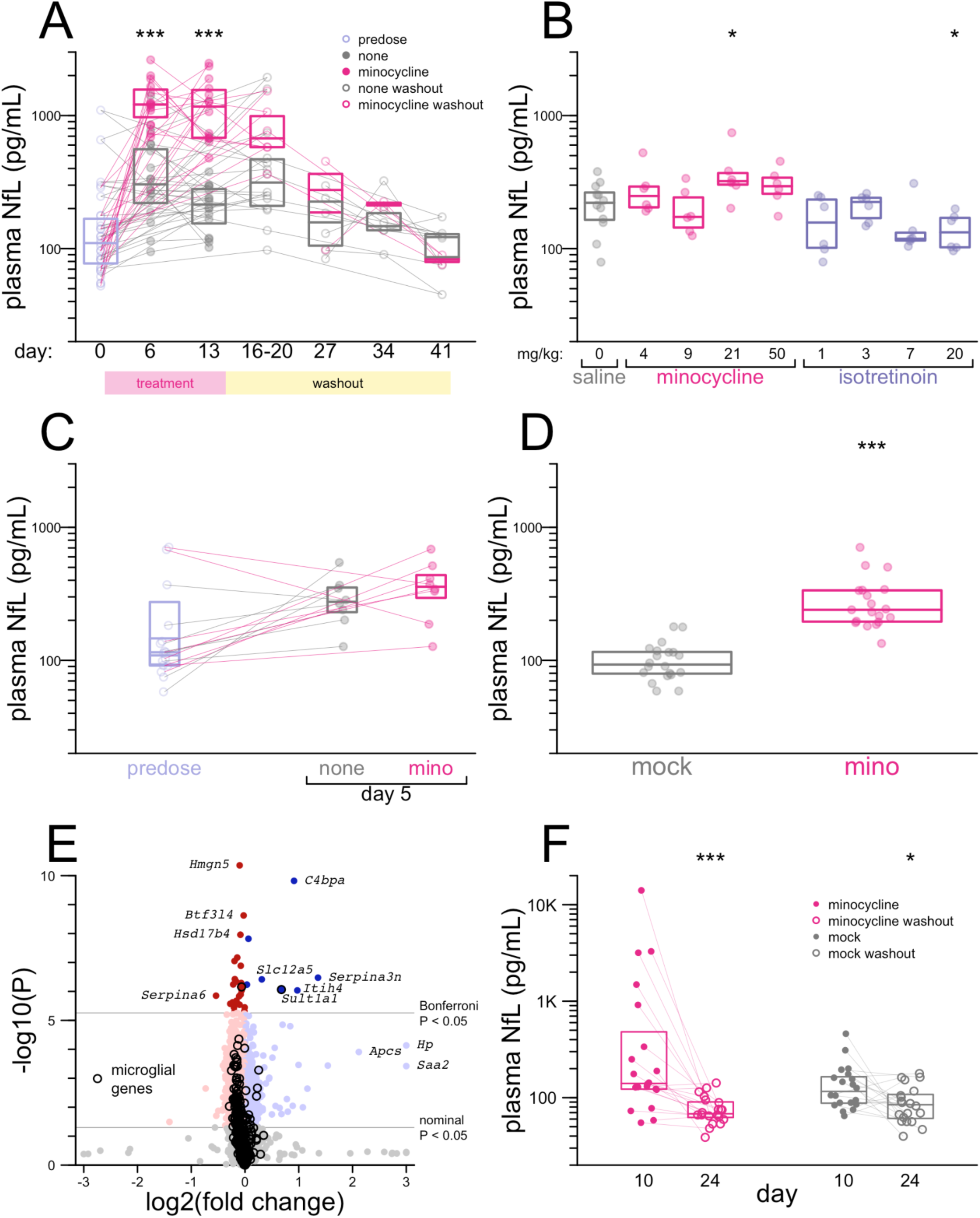
Neurofilament in mice treated with minocycline. **A)** Plasma NfL values in mouse study 1, with 50 mg/kg minocycline. See text for details. **B)** Plasma NfL values in mouse study 2, doses as indicated. **C)** Plasma NfL values in mouse study 3, with 50 mg/kg minocycline. **D)** Plasma NfL values in mouse study 4, after 5 days of 50 mg/kg minocycline. **E)** TMT on whole brain hemispheres of N=9 minocycline vs. N=9 mock-treated mice in mouse study 4, sacrificed at day 5. N=9,051 proteins quantified. **F)** Plasma NfL values in mouse study 4, after 10 days of 50 mg/kg minocycline and after 14 subsequent days of washout (no dosing). Key: * P < 0.05, ** P < 0.01, *** P < 0.001.

We sought to determine whether a better-tolerated dose of minocycline might also raise NfL. We also sought to determine the effect on plasma NfL of isotretinoin which, like minocycline, suppresses microglial response to LPS challenge in vivo^12^. We therefore conducted a 6-day, 4-point dose-response study for each compound with N=6 female animals per dose and N=12 in the saline group. In this study, the original 50 mg/kg dose of minocycline yielded only a non-significant 1.3-fold increase in plasma NfL compared to saline animals (median 294 vs. 221 pg/mL, P = 0.12, Wilcoxon test, Figure 4B, Table S18-S19). Isotretinoin did not increase NfL (Figure 4B). A repeat study of 5 days of 50 mg/kg dosing in N=8 mixed-sex animals per arm yielded similarly equivocal results (1.3-fold increase, median 358 vs. 276 pg/mL, P = 0.40, Wilcoxon test, Figure 4C, Table S20-S21).

Incorporating learnings from our study of the natural history of plasma NfL in mice (Figure 3), we conducted a fourth mouse study using N=20 female animals per group (Figure 4D, Table S22-S23). After 5 days of 50 mg/kg dosing, plasma NfL in minocycline-treated animals was 2.6-fold higher than untreated animals (median 240 vs. 93 pg/mL, P = 1.6e-7, Wilcoxon test, Figure 4D). Dosing for this duration was reasonably well-tolerated, although minocycline animals did lose 0.5% body weight on average over 5 days, a significant difference from the 2.4% weight gain in control animals (P = 4.4e-5, T test, Table S24-S25). Proteomic changes in the brains of mice that received minocycline for 5 days were relatively quiet (Figure 4E, Table S26). Microglial genes (Table S27) were disproportionately downregulated, but the vast majority were well below the significance threshold after multiple testing correction. Top significantly altered transcripts included complement-binding protein (*C4bpa*), and a microglia-expressed sulfotransferase involved in xenobiotic metabolism (*Sult1a1*) but did not collectively point to any one obvious pathway. Parenchymal expression of neurofilament itself was not increased. Markers of neuroinflammation (e.g. *Gfap*) and neurological insult (e.g. *Mapt*) were unaltered. In mice that received 10 days of minocycline followed by 14 days without dosing, median NfL dropped by 51% (from 140.5 to 62.8 pg/mL, P = 9.3e-5, Wilcoxon test), although a 27% decrease was also observed in mock-treated animals (84.6 vs. 116.0 pg/mL, P = 0.016, Wilcoxon test, Figure 4F). Overall, differences in magnitude of effect across the four mouse studies could not be readily attributed to any one experimental variable (Table S28).

We also sought to test whether minocycline might impact NfL concentration in conditioned media in a neuron-microglia co-culture system. After 65 days of differentiating cortical neurons from iPSC and 10 days of differentiating microglia into this system (see Methods), we began treatment with 25 µM minocycline or no compound. Conditioned media was completely removed every 2 days and subjected to NfL measurements, while new media was added. After 6 days of minocycline treatment, NfL increased by 2.0-fold from the baseline at day 0 (P = 0.065 vs. control wells). After minocycline was withdrawn, NfL continued to rise, reaching a 3.0-fold increase at day 10 (4 days post-withdrawal; P = 0.0043 vs. control wells; Figure 5A, Table S29-S30). Imaging of the co-cultures revealed no visually obvious difference in health between control and minocycline-treated cells (Figure 5B-C, Figure S3).

**Figure 5.**
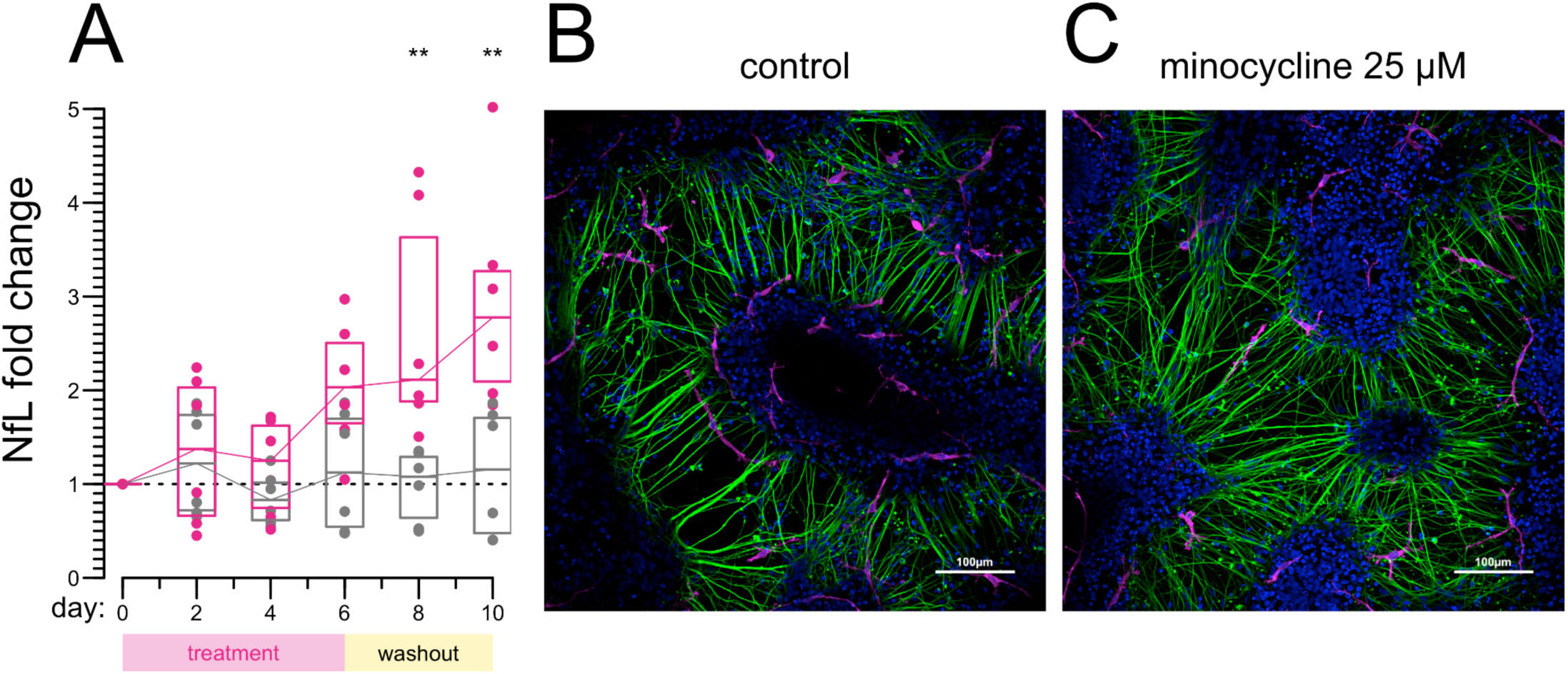
Neurofilament in conditioned media of neuron-microglia co-cultures. **A)** NfL concentration in conditioned media of neuron-microglia co-cultures treated with 25 µM minocycline or no treatment. Individual dots represent the N=6 biological replicates per condition, followed serially over 10 days; every 2 days, media was removed and analyzed for NfL, and new media was added. Boxes represent medians and interquartile ranges. ** P < 0.01 by Wilcoxon test. Raw and summarized values are provided in Tables S29-S30. **B-C)** Representative images of control (B) and minocycline-treated (C) co-cultures. Blue: DAPI; Green: Tuj1; Magenta: Iba1. Separate Tuj1 and Iba1 channels are provided in Figure S3.

## Discussion

Increases in NfL are generally assumed to reflect neuronal distress or damage. Prior studies where randomization to minocycline resulted in an increase in plasma NfL were interpreted as suggesting that minocycline exacerbated neurodegeneration, or was to some degree intrinsically neurotoxic^14, 17^. Correspondingly, low NfL has been interpreted as a neuroprotective signal^32^. Several observations here suggest that minocycline may be capable of modulating NfL via a neuronal health-independent mechanism.

First, both CSF and plasma NfL increased dramatically after 6 weeks’ minocycline treatment in a healthy research study participant. We likewise observed, variably, increases in NfL in plasma of wild-type, unchallenged mice treated with minocycline, and in conditioned media of co-cultured neurons and microglia.

Our research participant harbored a genetic predisposition to prion disease, but other than NfL, showed no biomarker evidence of disease process. Both the magnitude of this individual’s NfL increase — far beyond levels ever observed in prodromal genetic prion disease^20, 22, 23^ and indeed, higher than many symptomatic genetic prion disease patients^20, 33^, and the lack of disease onset two years later, argue that this spike does not correspond to the apparently very brief prodromal window in genetic prion disease. This individual also carried a dermatologic diagnosis which was the indication for minocycline treatment, but our analysis of discarded plasma from dermatology patients suggests that dermatologic conditions themselves are not a likely cause of elevated NfL. This individual took a 200 mg/day dose of minocycline, the most commonly prescribed clinical dose. If minocycline were so neurotoxic at this dose as to cause the magnitude of NfL increase observed here, one might expect this neurotoxicity to exhibit outward phenotypic manifestations, inconsistent with the relatively clean safety record and widespread clinical use of minocycline.

Second, the increase in NfL was highly specific. Other than a slight perturbation in microglial markers TREM2 and AIF1 and a decrease in intermediate filament VIM, no proteins other than NfL and NfM were altered in this individual’s CSF. If minocycline caused or exacerbated neurodegeneration, one might expect CSF T-tau to increase, which it did not. Similarly, in mice with elevated plasma NfL after 5 days of minocycline treatment, we observed a relatively quiet proteomic profile in brain parenchyma.

Third, the kinetics of NfL washout were rapid. In our study participant, NfL had returned almost to normal levels by 5 weeks after minocycline cessation. In 2 different mouse studies, we observed complete or nearly complete return to normal plasma NfL within 2 weeks of withdrawing minocycline. Spikes in plasma NfL due to acute neuronal insults are reported to take more than 12 weeks to clear^14, 34, 35^.

At the same time, many of the data collected here are inconsistent. We observed large and significant increases in NfL in only 2 of 4 studies of minocycline treatment in mice. Despite using the same dose (50 mg/kg/day) over similar dosing duration (5-6 days) and generating directionally consistent results, the other two studies saw small and non-significant NfL increases. Plasma NfL levels were extremely high in a subset of hospitalized patients taking minocycline, but not in others, and overall, any difference in plasma NfL between on-drug and off-drug timepoints was non-significant. Most of these inpatients were on only short courses of minocycline, and we counted them as “on” drug from the first day of treatment. Likewise our comparison of the mouse studies is based on a timepoint following 5-6 days of treatment. In contrast, our research participant had been taking minocycline for 6 weeks, and increases in plasma NfL in TBI and MS-CIS were observed after 12 weeks. One possible explanation for these discordant findings is that if minocycline inhibits the clearance or catabolism of NfL, then NfL concentration might increase rapidly (<1 week) in humans or mice that are releasing NfL rapidly, while it might rise much more slowly in humans releasing NfL at lower rates. CSF NfL was only modestly increased in *Grn* KO mice, and was not increased in *Trem2* KO mice, but was dramatically increased in *Grn/Trem2* double KO mice^36^, consistent with a “two-hit” hypothesis where the effect of inhibiting microglial clearance of NfL is magnified when there is also a perturbation that increases the rate of NfL release. Our hypothesis would also be consistent with data from Alzheimer’s patients indicating an inverse correlation between microglial activation and NfL^32^.

Based on our results, we hypothesize that microglia play a role in clearing NfL, and that minocycline inhibits this clearance, leading to an increase in CSF and plasma NfL concentration without causing neurodegeneration. At this time, however, this is only a hypothesis. It is not possible to make definitive conclusions, because our study has very prominent limitations. We possess detailed data on only one human research participant. Our data on dermatology patients and inpatients were collected only opportunistically, leading to incomplete clinical pictures, inconsistent numbers of days on drug, and sporadic plasma sampling. There appear to be pre-analytical variables in our discarded plasma samples that we do not fully understand. We did not implement a pharmacokinetic assay to determine drug concentration in the human plasma samples but instead relied on medical records of prescriptions and drug administrations. The poor pharmacokinetics of minocycline in rodents compelled us to administer minocycline at a dose ∼16 times the typical clinical dose. This high dose was not tolerated for 10 days, and even caused slight but significant weight loss by 5 days. Although this dose was not obviously toxic at 5-6 days, a timepoint when we already observed elevated plasma NfL, there remains doubt as to the degree to which toxicity may have contributed to the findings. Moreover, the massive variability in plasma NfL in mice means that some of our mouse studies may have been underpowered, and we do not know how much a lack of power versus genuine heterogeneity explains our discordant results.

While the evidence here is very preliminary, the implications of this hypothesis could be important. Given the pivotal role that NfL has begun to play in neurological clinical trials, the possibility that drugs — and perhaps also genetic, environmental, or other factors — might be capable of altering NfL concentration by a means other than causing or alleviating neurological insult, merits consideration.

## Methods

### Ethical approvals

Human samples were collected through Mass General Brigham (MGB) Institutional Review Board (IRB) protocols 2015P000221, 2017P000214, and 2021P002185. Our prion cohort research participant provided written informed consent. Discarded plasma samples were collected under a waiver of consent approved by the MGB IRB. The 4-digit hexadecimal IDs shown in the Supplementary Tables and Supplementary Figures were randomly generated and are not known to anyone outside the study group. Animal studies were performed under Broad Institute IACUC protocol 0162-05-17 and Charles River I026.

### Discarded plasma samples

Discarded plasma samples were collected through the Crimson Core Facility at Brigham and Women’s Hospital (“Crimson Core”) from between September 2021 and March 2023. Monthly queries in Epic were used to update a list of patients age 18-89 meeting the following criteria in the prior 90-day period: 1) a dermatology encounter and a diagnosis of acne, rosacea, dermatitis, or atopic dermatitis / eczema and a prescription start for minocycline, isotretinoin, doxycycline, clindamycin, or azithromycin; or 2) an annual physical exam encounter; or 3) inpatients with an order for minocycline. The list of matching medical record numbers was then flagged for plasma pulls at the Crimson Core at the point of sample discard. Any lavender top K_2_-EDTA tube plasma sample from one of these individuals for which all clinical testing was completed and 48 hours had passed was then pulled for inclusion in this study and labeled with a de-identified ID. Samples spent up to 8 hours at room temperature and up to 2 days at 4°C before being frozen at −20°C for transient storage and then shipped on dry ice to the analyzing laboratory for storage at −80°C. For individuals for whom plasma samples were obtained, sex, age, medication history, problem list, and, where applicable, date of death, were queried. Each individual was assigned a “zero” date corresponding to the earliest date of any prescription start or blood sample collected. Sample dates, prescription start and end dates, and death dates were then normalized to this zero. For analysis purposes, based on the duration of NfL elevation observed in the index case, patients were considered to be “on” drug from the prescription start date through 7 days after prescription end date. For outpatient prescriptions with no end date specified, the prescription was considered active for 30 days. Upon thaw, plasma samples were re-centrifuged and supernatants were used for NfL analysis. Ages were binned in 5-year intervals. Keys linking samples to medical record numbers were destroyed and all identifiable data were deleted; as noted above, the 4-digit hexadecimal IDs shown in the Supplementary Tables and Supplementary Figures were randomly generated and are not known to anyone outside the study group.

### Biomarker assays

NfL was determined per manufacturer’s instructions using ProteinSimple Ella (Bio-Techne). Total tau was determined by ELISA (Fujirebio). RT-QuIC was performed using the standard IQ-CSF protocol^37^.

### Animals and drug treatment

All animals were wild-type C57BL/6N age 6-20 weeks at study start from Charles River Laboratories. Animals were housed in groups of 4 on a 12/12 light/dark cycle with food and water ad libitum. Bleeds in the natural history study (Figure 3) and in minocycline treatment studies 3-4 (Figure 4C-F) were performed submandibular, while those in minocycline treatment studies 1-2 (Figure 4A-B) were performed via tail vein. Minocycline HCl (Gold Bio M-890-1) was made fresh daily in sterile saline and stored in the dark until intraperitoneal (i.p.) administration. Detailed parameters of each mouse study are compared in Table S28.

### Human induced pluripotent stem cell (hiPSC)-derived cortical neurons and microglia

hiPSC-derived cortical neurons were derived from the commercially acquired human cell line WTSIi015-A (Sigma) according to a published protocol^38^. Neural progenitor cells, induction day ∼30, were thawed and cultured in laminin-coated wells in neuronal maintenance media (NMM) with media change every other day. Final seed-out was performed on day 48 when cells were passaged and seeded on laminin-coated wells at a density of 5e4 cells/cm^2^. Cells were cultured for another week in NMM before hiPSC-derived microglia were added for co-cultures. For the differentiation to hiPSC-derived microglia, the previously described protocol was followed^38^. Briefly, hiPSCs are cultured on Matrigel (Corning) and after about 1.5-2 weeks embryonic body formation was initiated. Embryonic bodies were then plated into a 6-well-plate in hematopoietic medium (HM). After about 3-4 weeks with weekly media changes, primitive macrophage precursors can be harvested. Cells are counted, centrifuged, and resuspended in co-culture-media (CoM). CoM is NMM with addition of the growth factors Il-34 and GM-CSF at a concentration of 100 ng/mL and 10 ng/mL respectively. About 2.5e4 cells/cm^2^ are added to neuronal cultures and differentiated to microglia for 10 days before incubation with minocycline is started.

### Drug treatment of cell cultures

Cells were cultured in CoM until neuronal differentiation day 65 and microglial differentiation day 10 before addition of 25 µM minocycline (Sigma, #M9511-25mg) to CoM. Minocycline was dissolved in phosphate buffered saline solution (DPBS, Gibco, #14190094) to a concentration of 5 mg/mL and stored in aliquots at −80°C until used for cell culture experiments. Minocycline treatment lasted for 6 days, followed by a wash-out period of 4 days. Media was collected at the start of minocycline treatment as well as with media change every other day until the end of the experiment.

### NfL quantification in conditioned media

Every 2 days, the full volume of conditioned media was removed, centrifuged at 400 x *g* at 6°C for 7 min, and supernatants were frozen at −80°C until analysis. NfL levels were measured at the Clinical Neurochemistry Laboratory at the University of Gothenburg, Sweden. After thawing, samples were prepared for measurement with HD-X ultra-sensitive single molecule array (Simoa) platform (Quanterix). NfL was measured using the Neurology 4-Plex Kit (Lot # 503864, Quanterix) following the manufacturer’s instructions.

### Immunohistochemistry and microscopy

For immunohistochemistry, cells were seeded in Ibidi-slides (Ibidi, #80806). At the end of the experiment, cells were washed with PBS and fixated with Histofix (Histolab, #01000) for 15 minutes. Cells were washed with Tris-buffered saline (TBS), permeabilized for 15 minutes in permeabilization buffer (0.3% Triton X-100 in TBS) and blocked with blocking buffer (5% goat serum in permeabilization buffer) for 1h at room temperature. Cells were then incubated in primary antibody solution for Iba1 (1:500 in blocking buffer, Cell Signaling, #17198), TUJ1 (1:500 in blocking buffer, Abcam, ab78078) over night at 4°C. After a further wash with TBS, secondary antibody solution (Alexa-Fluor 568 goat anti-rabbit, Alexa-Fluor 488 goat anti-mouse) was added for 1h at room temperature. After another wash with TBS, samples were incubated for 5 min with the cell nuclei stain DAPI in TBS (1:1000), followed by another wash with TBS. Finally, Ibidi mounting media (Ibidi, #50001) was added and cells were visualized using a Nikon A1 inverted confocal microscope. Pictures presented are z-stacks covering 4 µm, merged for maximum intensity using Fiji ImageJ^39^.

### Transcriptomics

In mouse study 1, 200 mg pieces of cortex were dissected from whole hemispheres frozen in RNAlater. Bulk RNA-seq was performed on cortex from 3 minocycline-treated animals euthanized with weight loss on day 13, versus 8 untreated control animals harvested on day 16. RNA sequencing was performed by the Broad Institute Genomics Platform (Tru-Seq Strand Specific Large Insert RNA Sequencing) targeting 50 million read pairs. Transcript quantification was performed using Salmon^40^ on Terra.bio.

### Sample processing for proteomics - CSF samples

Prior to proteomics analysis, CSF samples from the individual at risk for genetic prion disease were first pre-processed in a dedicated prion laboratory consistent with best practices^41, 42^. 500 µL of each CSF sample was transferred to a depletion column to remove abundant plasma proteins (High-Select Top14 Abundant Protein Midi Spin Columns, Pierce), inverted several times to resuspend the depletion resin, then incubated on a rotisserie for 30 minutes at room temperature. The unbound fraction was retrieved by snapping off the column bottom end-cap, placing each column in a clean 15-mL tube and spinning at 1000 *g* for 3 minutes. Sodium dodecyl sulfate (SDS) was added to a final concentration of 2% (v/v), and dithiothreitol (DTT) to a final concentration of 5mM. After incubation at 37°C for 30 minutes samples were cooled to room temperature, treated with iodoacetamide (IAA; to 15mM final), and incubated in the dark for 1 hour at room temperature. Alkylation was quenched by adding DTT to 10mM final concentration. A protein pellet was then isolated by methanol chloroform precipitation, air dried, denatured in 8 M guanidine hydrochloride (GdnHCl) for 30 minutes, then diluted with 100mM EPPS buffer pH 8.1. Digestion was then performed with LysC (Promega V1671 at 1:50 enzyme:protein) overnight at room temperature followed by trypsin (Promega V5113 at 1:50 enzyme:protein) for 37°C for 6 hours.

### Sample processing for proteomics - brain samples

Brains from uninfected mice were first homogenized at 10% wt/vol in 0.2% CHAPS as described^43^ and homogenates were frozen at −80°C. Upon thaw, homogenates were treated with 4 M urea + 1% SDS + 100 mM EPPS pH 8.0 + HALT protease and phosphatase inhibitors (Thermo Fisher Scientific) and bead beaten at 2°C using a Precellys Evolution homogenizer for a total of 3 cycles: 10 second intervals at 7200 rpm with 60 second pauses. Protein concentration in the lysates were quantified using the Pierce micro-BCA assay (Thermo Fisher Scientific). Protein reduction was performed for 60 min at 25°C with 5 mM dithiothreitol (DTT). Free thiol containing cysteines were alkylated with 15 mM iodoacetamide and quenched with further addition of DTT to a final 10 mM prior to protein purification. The single-pot, solid-phase-enhanced, sample preparation (SP3) protocol^44, 45^ was applied to purify protein across all samples in these studies. Briefly, the magnetic beads were prepared by combining E3 and E7 Sera-Mag Carboxylate-Modified Magnetic SpeedBeads (1:1; vol:vol) (Cytiva). The beads were washed three times with HPLC H2O prior to resuspension in lysis buffer (1% SDS, 100 mM EPPS, pH 8.0) for a final bead slurry concentration of 50 µg/µL. Approximately 20 µL of the pre-washed bead slurry and 75 µL 100% ethanol was added to each 30 µL sample containing 50 µg of protein (4M urea, 1% SDS, 100 mM EPPS pH 8.0). Proteins were allowed to bind and incubate with the bead slurry for 15 min at 25°C with periodic gentle vortexing. Following protein binding, the supernatant was discarded, and the bead-captured proteins were washed three times with 80% ethanol and air dried. The purified protein-captured beads were resuspended in 100 mM EPPS, pH 8.0. Approximately 50 µg of protein/sample was digested at 25°C for 12 hours with lysyl endopeptidase (LysC, Wako Chemicals USA) at a 1:12.5 (w/w) protease:protein ratio. Following LysC digestion, the peptides were digested with trypsin at 37 C for 8 hours (Promega) at a 1:25 (w/w) protease:protein ratio. The digested peptides (supernatant) were purified and separated from the magnetic beads. Three post-digestion washes were performed to wash and ensure elution of all peptides from the beads with the following buffers: 100 mM EPPS pH 8.0, 0.2% formic acid, and 80% acetonitrile/0.1% formic acid. With every wash and elution, the supernatant was transferred and combined with the initial supernatant and dried to completion by vacuum centrifugation. The percentage of missed cleavages was monitored across 4 random samples to ensure optimal digestion efficiency prior to isobaric labeling.

### TMT labeling and fractionation - CSF samples

Following protein digestion with LysC and trypsin, the percentage of missed cleavages was monitored across four random samples, desalted, and analyzed by mass spectrometry to ensure optimal digestion efficiency prior to isobaric labeling. Isobaric labeling of peptides was performed with the 10-plex tandem mass tag (TMT) reagents (Thermo Fisher Scientific). The TMT10 reagents (120 µg per channel) were added to all samples and incubated for 2 hours at 25° C. A small portion (5%) of each sample was mixed together, desalted via Empore-C18 StageTip, and analyzed via LC-MS to check TMT labeling efficiency and loading ratio. All labeling reactions were quenched with hydroxylamine (0.5% final) and acidified with trifluoroacetic acid (2% final). The ten samples were mixed according to total summed TMT signal observed in the labeling efficiency check such that all samples have equal loading. The pooled TMT10 labeled peptide mix was desalted with a 50 mg tC18 Sep-Pak (Waters) and dried by vacuum centrifugation. The desalted, pooled TMT labeled peptides were fractionated using a high pH reverse-phase peptide fractionation kit (Pierce) into seven total fractions (10%, 15%, 17.5%, 20%, 30%, 50% and 70% acetonitrile in 0.1% triethylamine) and vacuum dried. The fractions were desalted on C18 StageTips and analyzed using a 180min SPS-MS3 method on an Orbitrap Eclipse instrument.

### TMT labeling and fractionation - brain samples

All peptides (50 µg/sample) were resolubilized in 20 µL 500 mM EPPS, pH 8.0. Isobaric labeling of peptides was performed with the 18-plex tandem mass tag (TMT) reagents (Thermo Fisher Scientific). The TMTPro reagents (5 mg) were resuspended in 100 µL anhydrous acetonitrile (ACN) and 5 µL (200 µg) was added to all of the peptides and incubated for 2 hours at 25° C. A small portion (1%) of each sample was mixed together, desalted via C18 StageTip, and analyzed via LC-MS to check TMT labeling efficiency and loading ratio. All labeling reactions were quenched with hydroxylamine (0.5% final) and acidified with trifluoroacetic acid (2% final). The 18 channels were mixed according to total summed TMT signal observed in the labeling efficiency check such that all samples have equal loading. The final TMT mixed sample was desalted with a 50 mg tC18 Sep-Pak (Waters) and dried by vacuum centrifugation. Approximately 100 µg of peptide mix was subjected to orthogonal basic-pH reverse phase fractionation on a 3×150 mm column packed with 1.9 µm Poroshell C18 material (Agilent, Santa Clara, CA), utilizing a 45 min linear gradient from 86% buffer A (5% acetonitrile in 10 mM ammonium bicarbonate, pH 8) to 42% buffer B (90% acetonitrile in 10 mM ammonium bicarbonate, pH 8) at a flow rate of 0.3 ml/min. Ninety six fractions were collected and consolidated into 24 total fractions, acidified with formic acid and vacuum dried. The fractions were resuspended in 0.2% formic acid, desalted on StageTips and vacuum dried. Peptides were reconstituted in 5% formic acid + 5% acetonitrile for LC-MS3 analysis.

### Mass spectrometry analysis

All mass spectra were acquired on an Orbitrap instrument (Eclipse for CSF samples, Lumos for brain) coupled to an EASY nanoLC-1200 (Thermo Fisher) liquid chromatography system. Peptides (approximately 1 µg for CSF, 2 µg for brain) were loaded on a 75 µm capillary column packed in-house with Sepax GP-C18 resin (1.8 µm, 150 Å, Sepax) to a final length of 40 cm (CSF) or 35 cm (brain). Peptides for total protein analysis were separated using a 180 minute (CSF) or 90 minute (brain) linear gradient from 15% (CSF) or 14% (brain) to 40% acetonitrile in 0.1% formic acid. The mass spectrometer was operated in a data dependent mode. The scan sequence began with FTMS1 spectra (resolution = 120,000; mass range of 350-1400 *m/z*; max injection time of 10 ms; AGC target of 5e5; dynamic exclusion for 75 seconds (CSF) or 60 seconds (brain) with a +/- 10 ppm window). The most intense precursor ions were selected for ITMS2 analysis via collisional-induced dissociation (CID) in the ion trap (normalized collision energy (NCE) = 35; max injection time = 35ms; isolation window of 0.7 Da; AGC target of 1e4) within a 2 second window. A real-time search approach^46^ was utilized during data acquisition to only trigger quantitative spectra on high-confidence peptide identifications. Online spectral identification was accomplished via a custom software client that monitors spectral acquisition though a vendor supplied instrument application programming interface and assigns peptide sequences through a probabilistic model, in real-time. The RTS client utilized a peptide database composed of *in-silico* predicted species-specific tryptic peptides. All mass spectra were converted to mzXML using a modified version of ReAdW.exe. MS/MS spectra were searched against a concatenated Uniprot species-specific protein database containing common contaminants (forward + reverse sequences) using the SEQUEST algorithm^47^. Database search criteria are as follows: fully tryptic with two missed cleavages; a precursor mass tolerance of 50 ppm and a fragment ion tolerance of 1 Da for peptides; oxidation of methionine (15.9949 Da) was set as a differential modification. Static modifications were iodoacetamide on cysteines (57.02146) and TMT on lysines and N-termini of peptides (CSF: TMT10: 229.1629 Da; brain: TMT18: 304.2071). Peptide-spectrum matches were filtered using linear discriminant analysis^48^ and adjusted to a 1% peptide false discovery rate (FDR)^49^ and collapsed further to a final 1.0% protein-level FDR. Proteins were quantified by summing the total reporter intensities across all matching PSMs.

### Statistics, data, and source code availability

All analyses were performed in R 4.2.0. Because NfL concentrations are non-normally distributed, they were visualized using medians and interquartile ranges and were compared using Wilcoxon tests. The dermatology cohort was analyzed using log-linear regression: lm(log(nfl) ∼ drug + age). Because some inpatients had both “on” and “off” samples while others had both, inpatient data were compared using a partially paired Wilcoxon implemented in the R package robustrank. The volcano plot of TMT data in minocycline-treated mice was produced by grouping peptides by gene symbol using Empirical Brown’s Method^50^ (R Bioconductor) with subsequent Bonferroni correction. P values less than 0.05 were considered nominally significant. Transcriptomic analysis used DESeq2^51^ on Terra (Terra.bio) with default parameters and Benjamini-Hochberg control of false discovery rate. Genes were annotated as microglial if they had mean count >0.1 in microglia and log2 fold change >2 with nominal P < 0.05 compared to all other cell types based on mouse cortex 10X single cell data^52^ analyzed in Loupe Browser. Source code and an analytical dataset sufficient to reproduce all figures and statistics in this manuscript will be made publicly available at https://github.com/ericminikel/mino

## Data Availability

Source code and an analytical dataset sufficient to reproduce all figures and statistics in this manuscript will be made publicly available at https://github.com/ericminikel/mino

## Acknowledgments

We thank all research participants for their contributions to this study. This work was funded by Prion Alliance, the Broad Institute (BroadIgnite Accelerator), donations to the Prions@Broad fund, and the National Institutes of Health (R21 TR003040, R01 NS125255, R01 NS132022).

## SUPPLEMENT

Supplementary Tables are provided as a separate Excel file.

**Figure S1.**
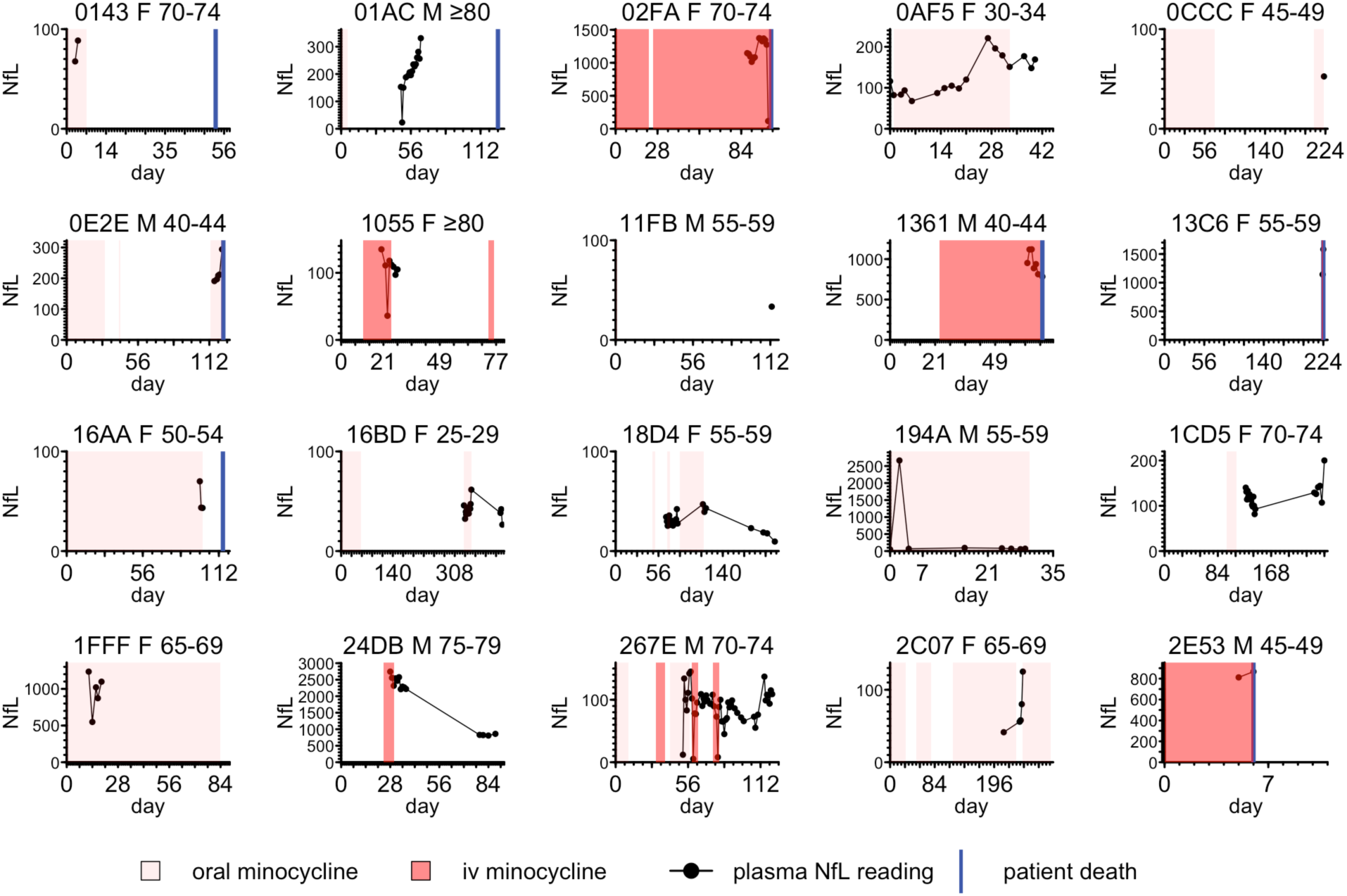
Individual inpatient NfL trajectories and minocycline treatment. Shown above each panel is a de-identified patient ID, which matches diagnosis and drug indication details shown in Table S6, and the patient’s sex and age. As noted in Methods, the 4-digit hexadecimal IDs used above were randomly generated and are not known to anyone outside the study group.

**Figure S2.**
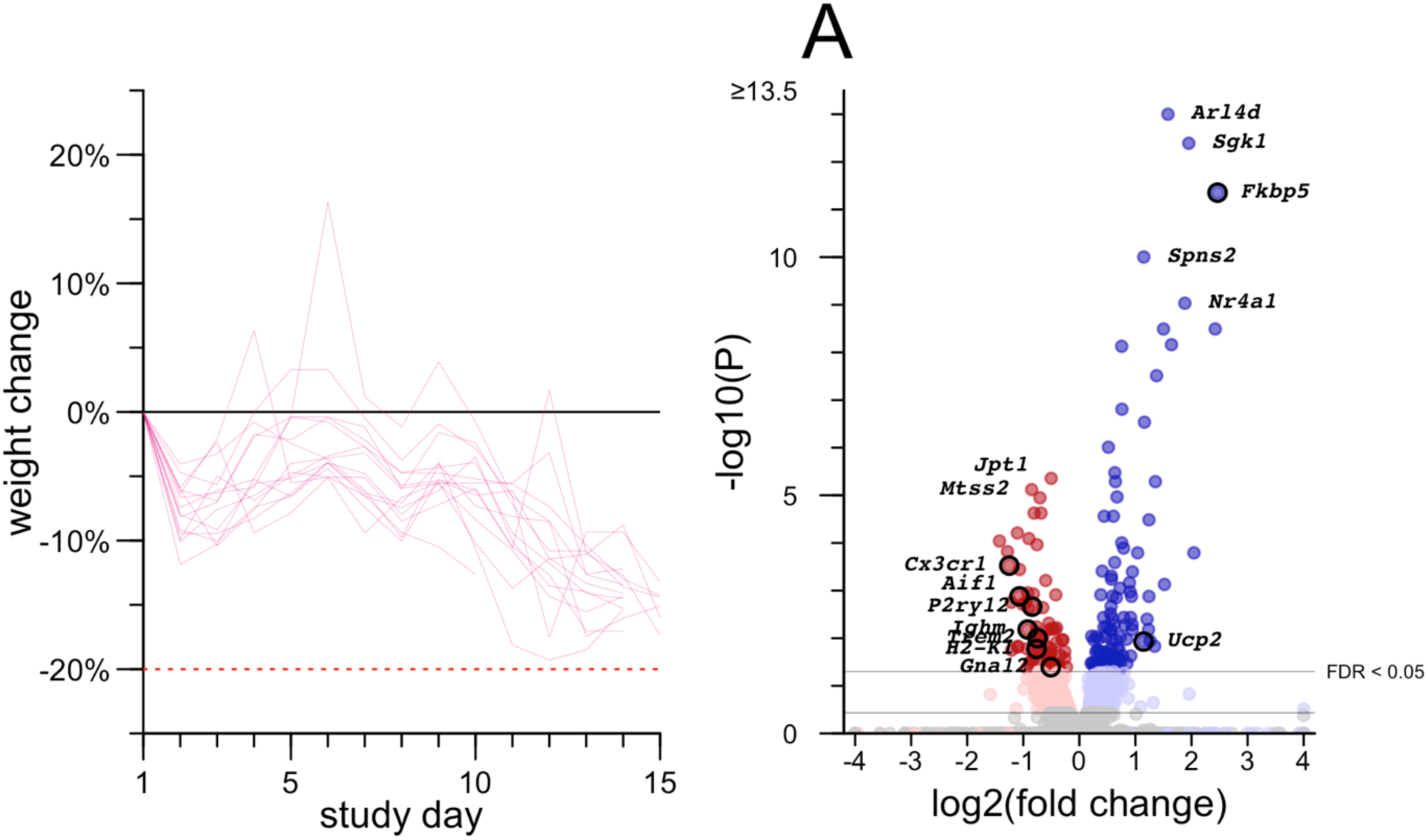
Additional results from mouse study 1. Left: individual weight trajectories for mice treated with 50 mg/kg/day minocycline. Right: volcano plot of differentially expressed genes in the brains of these mice by bulk RNA-seq analyzed with DESeq2, see Methods for details.

**Figure S3.**
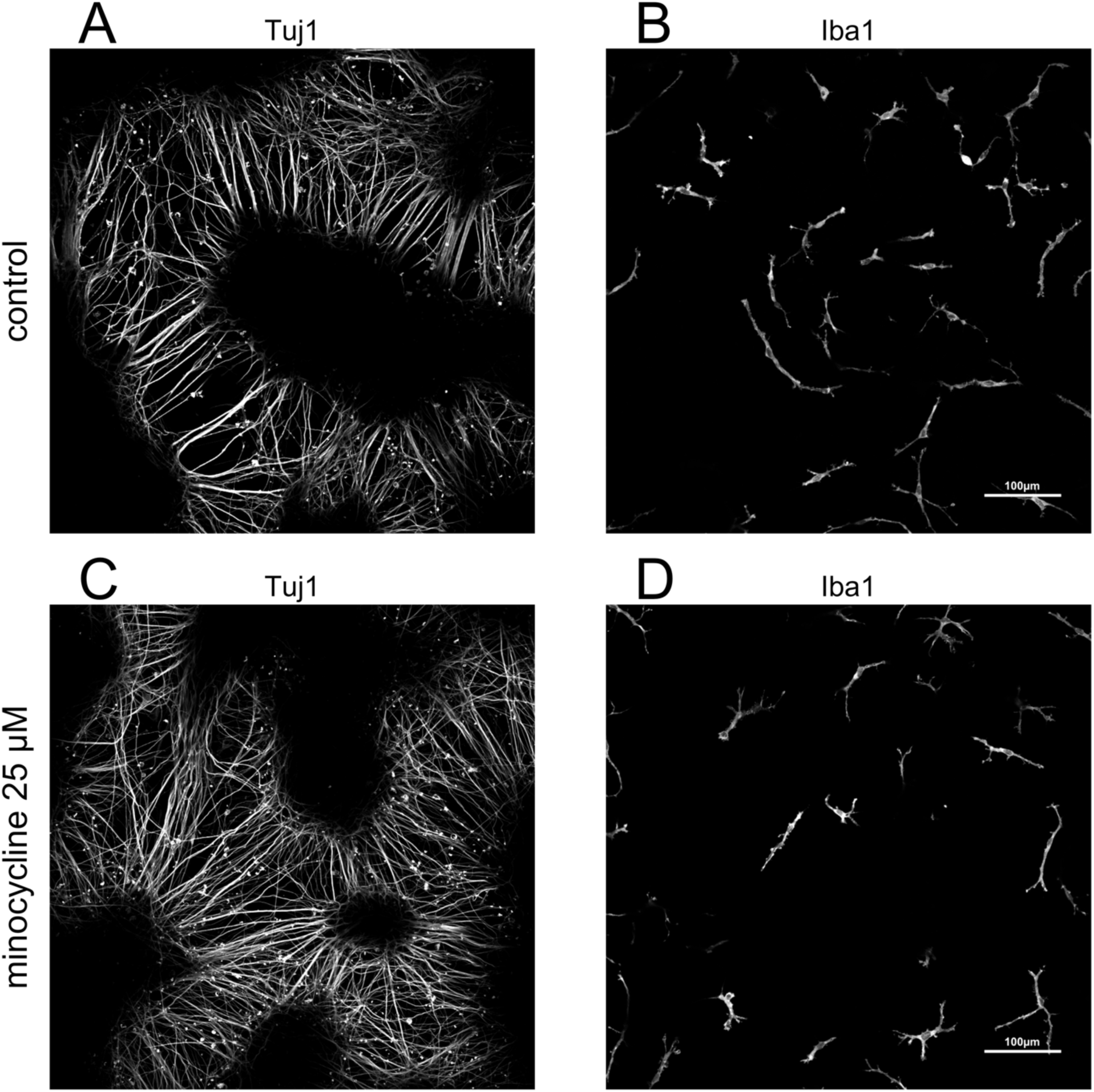
Co-culture images broken out by color channel. Control (A-B) and minocycline (C-D) images from Figure 5, broken out by green Tuj1 channel (A, C) and magenta Iba1 channel (B, D).

## References

1. Yuan A, Rao MV, Veeranna null, Nixon RA. Neurofilaments and Neurofilament Proteins in Health and Disease. Cold Spring Harb Perspect Biol. 2017 Apr 3;9(4):a018309. PMCID: PMC5378049

2. Bridel C, van Wieringen WN, Zetterberg H, Tijms BM, Teunissen CE, and the NFL Group, Alvarez-Cermeño JC, Andreasson U, Axelsson M, Bäckström DC, Bartos A, Bjerke M, Blennow K, Boxer A, Brundin L, Burman J, Christensen T, Fialová L, Forsgren L, Frederiksen JL, Gisslén M, Gray E, Gunnarsson M, Hall S, Hansson O, Herbert MK, Jakobsson J, Jessen-Krut J, Janelidze S, Johannsson G, Jonsson M, Kappos L, Khademi M, Khalil M, Kuhle J, Landén M, Leinonen V, Logroscino G, Lu CH, Lycke J, Magdalinou NK, Malaspina A, Mattsson N, Meeter LH, Mehta SR, Modvig S, Olsson T, Paterson RW, Pérez-Santiago J, Piehl F, Pijnenburg YAL, Pyykkö OT, Ragnarsson O, Rojas JC, Romme Christensen J, Sandberg L, Scherling CS, Schott JM, Sellebjerg FT, Simone IL, Skillbäck T, Stilund M, Sundström P, Svenningsson A, Tortelli R, Tortorella C, Trentini A, Troiano M, Turner MR, van Swieten JC, Vågberg M, Verbeek MM, Villar LM, Visser PJ, Wallin A, Weiss A, Wikkelsø C, Wild EJ. Diagnostic Value of Cerebrospinal Fluid Neurofilament Light Protein in Neurology: A Systematic Review and Meta-analysis. JAMA Neurol. 2019 Sep 1;76(9):1035–1048. PMCID: PMC6580449

3. Ashton NJ, Janelidze S, Al Khleifat A, Leuzy A, van der Ende EL, Karikari TK, Benedet AL, Pascoal TA, Lleó A, Parnetti L, Galimberti D, Bonanni L, Pilotto A, Padovani A, Lycke J, Novakova L, Axelsson M, Velayudhan L, Rabinovici GD, Miller B, Pariante C, Nikkheslat N, Resnick SM, Thambisetty M, Schöll M, Fernández-Eulate G, Gil-Bea FJ, López de Munain A, Al-Chalabi A, Rosa-Neto P, Strydom A, Svenningsson P, Stomrud E, Santillo A, Aarsland D, van Swieten JC, Palmqvist S, Zetterberg H, Blennow K, Hye A, Hansson O. A multicentre validation study of the diagnostic value of plasma neurofilament light. Nat Commun. 2021 Jun 7;12(1):3400. PMCID: PMC8185001

4. Kuhle J, Kropshofer H, Haering DA, Kundu U, Meinert R, Barro C, Dahlke F, Tomic D, Leppert D, Kappos L. Blood neurofilament light chain as a biomarker of MS disease activity and treatment response. Neurology. 2019 Mar 5;92(10):e1007–e1015. PMCID: PMC6442011

5. van Roon-Mom W, Ferguson C, Aartsma-Rus A. From Failure to Meet the Clinical Endpoint to U.S. Food and Drug Administration Approval: 15th Antisense Oligonucleotide Therapy Approved Qalsody (Tofersen) for Treatment of SOD1 Mutated Amyotrophic Lateral Sclerosis. Nucleic Acid Ther. 2023 Aug;33(4):234–237. PMID: 37581487

6. Benatar M, Wuu J, Andersen PM, Andrews J, Bucelli RC, Otto M, Ferguson TA, Chen W, Fanning L, Graham D, Sun P, Liu Y, Wong J, Fradette S. Design of a Phase 3, Randomized, Placebo-controlled Trial of Tofersen Initiated in Clinically Pre-symptomatic SOD1 Mutation Carriers with a Longitudinal Natural History Run-in (2285). Neurology. 2021 Apr 13;96(15 Supplement):2285.

7. Krach F, Stemick J, Boerstler T, Weiss A, Lingos I, Reischl S, Meixner H, Ploetz S, Farrell M, Hehr U, Kohl Z, Winner B, Winkler J. An alternative splicing modulator decreases mutant HTT and improves the molecular fingerprint in Huntington’s disease patient neurons. Nat Commun. 2022 Nov 10;13(1):6797. PMCID: PMC9649613

8. Harding R. Disappointing news from Novartis about branaplam and the VIBRANT-HD trial [Internet]. HDBuzz. 2022 [cited 2024 Apr 4]. Available from: https://en.hdbuzz.net/338

9. Asadi A, Abdi M, Kouhsari E, Panahi P, Sholeh M, Sadeghifard N, Amiriani T, Ahmadi A, Maleki A, Gholami M. Minocycline, focus on mechanisms of resistance, antibacterial activity, and clinical effectiveness: Back to the future. J Glob Antimicrob Resist. 2020 Sep;22:161–174. PMID: 32061815

10. Barbieri JS, Bhate K, Hartnett KP, Fleming-Dutra KE, Margolis DJ. Trends in Oral Antibiotic Prescription in Dermatology, 2008 to 2016. JAMA Dermatol. 2019 Mar 1;155(3):290–297. PMCID: PMC6439939

11. Garner SE, Eady A, Bennett C, Newton JN, Thomas K, Popescu CM. Minocycline for acne vulgaris: efficacy and safety. Cochrane Database Syst Rev. 2012 Aug 15;2012(8):CD002086. PMCID: PMC7017847

12. Shankaran M, Marino ME, Busch R, Keim C, King C, Lee J, Killion S, Awada M, Hellerstein MK. Measurement of brain microglial proliferation rates in vivo in response to neuroinflammatory stimuli: application to drug discovery. J Neurosci Res. 2007 Aug 15;85(11):2374–2384. PMID: 17551981

13. Schafer DP, Lehrman EK, Kautzman AG, Koyama R, Mardinly AR, Yamasaki R, Ransohoff RM, Greenberg ME, Barres BA, Stevens B. Microglia sculpt postnatal neural circuits in an activity and complement-dependent manner. Neuron. 2012 May 24;74(4):691–705. PMCID: PMC3528177

14. Scott G, Zetterberg H, Jolly A, Cole JH, De Simoni S, Jenkins PO, Feeney C, Owen DR, Lingford-Hughes A, Howes O, Patel MC, Goldstone AP, Gunn RN, Blennow K, Matthews PM, Sharp DJ. Minocycline reduces chronic microglial activation after brain trauma but increases neurodegeneration. Brain. 2018 Feb 1;141(2):459–471. PMCID: PMC5837493

15. Panizzutti B, Skvarc D, Lin S, Croce S, Meehan A, Bortolasci CC, Marx W, Walker AJ, Hasebe K, Kavanagh BE, Morris MJ, Mohebbi M, Turner A, Gray L, Berk L, Walder K, Berk M, Dean OM. Minocycline as Treatment for Psychiatric and Neurological Conditions: A Systematic Review and Meta-Analysis. Int J Mol Sci. 2023 Mar 9;24(6):5250. PMCID: PMC10049047

16. Gordon PH, Moore DH, Miller RG, Florence JM, Verheijde JL, Doorish C, Hilton JF, Spitalny GM, MacArthur RB, Mitsumoto H, Neville HE, Boylan K, Mozaffar T, Belsh JM, Ravits J, Bedlack RS, Graves MC, McCluskey LF, Barohn RJ, Tandan R, Western ALS Study Group. Efficacy of minocycline in patients with amyotrophic lateral sclerosis: a phase III randomised trial. Lancet Neurol. 2007 Dec;6(12):1045– 1053. PMID: 17980667

17. Camara-Lemarroy C, Metz L, Kuhle J, Leppert D, Willemse E, Li DK, Traboulsee A, Greenfield J, Cerchiaro G, Silva C, Yong VW. Minocycline treatment in clinically isolated syndrome and serum NfL, GFAP, and metalloproteinase levels. Mult Scler. 2022 Nov;28(13):2081–2089. PMCID: PMC9574233

18. Abu-Rumeileh S, Capellari S, Stanzani-Maserati M, Polischi B, Martinelli P, Caroppo P, Ladogana A, Parchi P. The CSF neurofilament light signature in rapidly progressive neurodegenerative dementias. Alzheimers Res Ther. 2018 Jan 11;10(1):3. PMCID: PMC5784714

19. Zerr I, Schmitz M, Karch A, Villar-Piqué A, Kanata E, Golanska E, Díaz-Lucena D, Karsanidou A, Hermann P, Knipper T, Goebel S, Varges D, Sklaviadis T, Sikorska B, Liberski PP, Santana I, Ferrer I, Zetterberg H, Blennow K, Calero O, Calero M, Ladogana A, Sánchez-Valle R, Baldeiras I, Llorens F. Cerebrospinal fluid neurofilament light levels in neurodegenerative dementia: Evaluation of diagnostic accuracy in the differential diagnosis of prion diseases. Alzheimers Dement. 2018 Jun;14(6):751–763. PMID: 29391125

20. Thompson AGB, Anastasiadis P, Druyeh R, Whitworth I, Nayak A, Nihat A, Mok TH, Rudge P, Wadsworth JDF, Rohrer J, Schott JM, Heslegrave A, Zetterberg H, Collinge J, Jackson GS, Mead S. Evaluation of plasma tau and neurofilament light chain biomarkers in a 12-year clinical cohort of human prion diseases. Mol Psychiatry. 2021 Mar 5; PMID: 33674752

21. Vallabh SM, Minikel EV, Williams VJ, Carlyle BC, McManus AJ, Wennick CD, Bolling A, Trombetta BA, Urick D, Nobuhara CK, Gerber J, Duddy H, Lachmann I, Stehmann C, Collins SJ, Blennow K, Zetterberg H, Arnold SE. Cerebrospinal fluid and plasma biomarkers in individuals at risk for genetic prion disease. BMC Med. 2020 Jun 18;18(1):140. PMCID: PMC7302371

22. Mok TH, Nihat A, Majbour N, Sequeira D, Holm-Mercer L, Coysh T, Darwent L, Batchelor M, Groveman BR, Orrù CD, Hughson AG, Heslegrave A, Laban R, Veleva E, Paterson RW, Keshavan A, Schott J, Swift IJ, Heller C, Rohrer JD, Gerhard A, Butler C, Rowe JB, Masellis M, Chapman M, Lunn MP, Bieschke J, Jackson GS, Zetterberg H, Caughey B, Rudge P, Collinge J, Mead S. Seed amplification and neurodegeneration marker trajectories in individuals at risk of prion disease. Brain. 2023 Mar 28;awad101. PMID: 36975162

23. Sonia M Vallabh, Meredith A Mortberg, Shona W Allen, Ashley C Kupferschmid, Pia Kivisakk, Bruno L Hammerschlag, Anna Bolling, Bianca A Trombetta, Kelli Devitte-McKee, Abaigeal M Ford, Lauren Sather, Griffin Duffy, Ashley Rivera, Jessica Gerber, Alison J McManus, Eric Vallabh Minikel, Steven E Arnold. Biomarker changes preceding symptom onset in genetic prion disease. medRxiv. 2023 Dec 18;2023.12.18.23300042.

24. Minikel EV, Vallabh SM. Where have prions been all our lives? Brain. 2023 May 10;awad143. PMID: 37161596

25. Bornhorst JA, Figdore D, Campbell MR, Pazdernik VK, Mielke MM, Petersen RC, Algeciras-Schimnich A. Plasma neurofilament light chain (NfL) reference interval determination in an Age-stratified cognitively unimpaired cohort. Clin Chim Acta. 2022 Oct 1;535:153–156. PMID: 36041549

26. Hviid CVB, Knudsen CS, Parkner T. Reference interval and preanalytical properties of serum neurofilament light chain in Scandinavian adults. Scand J Clin Lab Invest. 2020 Jul;80(4):291–295. PMID: 32077769

27. Simrén J, Ashton NJ, Blennow K, Zetterberg H. Blood neurofilament light in remote settings: Alternative protocols to support sample collection in challenging pre-analytical conditions. Alzheimers Dement (Amst). 2021;13(1):e12145. PMCID: PMC7896630

28. Disanto G, Barro C, Benkert P, Naegelin Y, Schädelin S, Giardiello A, Zecca C, Blennow K, Zetterberg H, Leppert D, Kappos L, Gobbi C, Kuhle J, Swiss Multiple Sclerosis Cohort Study Group. Serum Neurofilament light: A biomarker of neuronal damage in multiple sclerosis. Ann Neurol. 2017 Jun;81(6):857–870. PMCID: PMC5519945

29. Agwuh KN, MacGowan A. Pharmacokinetics and pharmacodynamics of the tetracyclines including glycylcyclines. J Antimicrob Chemother. 2006 Aug;58(2):256–265. PMID: 16816396

30. Bowers DR, Cao H, Zhou J, Ledesma KR, Sun D, Lomovskaya O, Tam VH. Assessment of minocycline and polymyxin B combination against Acinetobacter baumannii. Antimicrob Agents Chemother. 2015 May;59(5):2720–2725. PMCID: PMC4394818

31. Zhou J, Ledesma KR, Chang KT, Abodakpi H, Gao S, Tam VH. Pharmacokinetics and Pharmacodynamics of Minocycline against Acinetobacter baumannii in a Neutropenic Murine Pneumonia Model. Antimicrob Agents Chemother. 2017 May;61(5):e02371–16. PMCID: PMC5404596

32. Parbo P, Madsen LS, Ismail R, Zetterberg H, Blennow K, Eskildsen SF, Vorup-Jensen T, Brooks DJ. Low plasma neurofilament light levels associated with raised cortical microglial activation suggest inflammation acts to protect prodromal Alzheimer’s disease. Alzheimers Res Ther. 2020 Jan 2;12(1):3. PMCID: PMC6941285

33. Abu-Rumeileh S, Baiardi S, Ladogana A, Zenesini C, Bartoletti-Stella A, Poleggi A, Mammana A, Polischi B, Pocchiari M, Capellari S, Parchi P. Comparison between plasma and cerebrospinal fluid biomarkers for the early diagnosis and association with survival in prion disease. J Neurol Neurosurg Psychiatry. 2020 Sep 14; PMID: 32928934

34. Shahim P, Zetterberg H, Tegner Y, Blennow K. Serum neurofilament light as a biomarker for mild traumatic brain injury in contact sports. Neurology. 2017 May 9;88(19):1788–1794. PMCID: PMC5419986

35. Shahim P, Politis A, van der Merwe A, Moore B, Chou YY, Pham DL, Butman JA, Diaz-Arrastia R, Gill JM, Brody DL, Zetterberg H, Blennow K, Chan L. Neurofilament light as a biomarker in traumatic brain injury. Neurology. 2020 Aug 11;95(6):e610–e622. PMCID: PMC7455357

36. Reifschneider A, Robinson S, van Lengerich B, Gnörich J, Logan T, Heindl S, Vogt MA, Weidinger E, Riedl L, Wind K, Zatcepin A, Pesämaa I, Haberl S, Nuscher B, Kleinberger G, Klimmt J, Götzl JK, Liesz A, Bürger K, Brendel M, Levin J, Diehl-Schmid J, Suh J, Di Paolo G, Lewcock JW, Monroe KM, Paquet D, Capell A, Haass C. Loss of TREM2 rescues hyperactivation of microglia, but not lysosomal deficits and neurotoxicity in models of progranulin deficiency. EMBO J. 2022 Feb 15;41(4):e109108. PMCID: PMC8844989

37. Orrú CD, Groveman BR, Hughson AG, Zanusso G, Coulthart MB, Caughey B. Rapid and sensitive RT-QuIC detection of human Creutzfeldt-Jakob disease using cerebrospinal fluid. MBio. 2015;6(1). PMCID: PMC4313917

38. Fruhwürth S, Reinert LS, Öberg C, Sakr M, Henricsson M, Zetterberg H, Paludan SR. TREM2 is down-regulated by HSV1 in microglia and involved in antiviral defense in the brain. Sci Adv. 2023 Aug 18;9(33):eadf5808. PMCID: PMC10438464

39. Schindelin J, Arganda-Carreras I, Frise E, Kaynig V, Longair M, Pietzsch T, Preibisch S, Rueden C, Saalfeld S, Schmid B, Tinevez JY, White DJ, Hartenstein V, Eliceiri K, Tomancak P, Cardona A. Fiji: an open-source platform for biological-image analysis. Nat Methods. 2012 Jun 28;9(7):676–682. PMCID: PMC3855844

40. Patro R, Duggal G, Love MI, Irizarry RA, Kingsford C. Salmon provides fast and bias-aware quantification of transcript expression. Nat Methods. 2017 Apr;14(4):417–419. PMCID: PMC5600148

41. Moore RA, Ward A, Race B, Priola SA. Processing of high-titer prions for mass spectrometry inactivates prion infectivity. Biochim Biophys Acta Proteins Proteom. 2018;1866(11):1174–1180. PMID: 30282615

42. Minikel EV, Kuhn E, Cocco AR, Vallabh SM, Hartigan CR, Reidenbach AG, Safar JG, Raymond GJ, McCarthy MD, O’Keefe R, Llorens F, Zerr I, Capellari S, Parchi P, Schreiber SL, Carr SA. Domain-specific quantification of prion protein in cerebrospinal fluid by targeted mass spectrometry. Mol Cell Proteomics. 2019 Sep 26; PMID: 31558565

43. Mortberg MA, Zhao HT, Reidenbach AG, Gentile JE, Kuhn E, O’Moore J, Dooley PM, Connors TR, Mazur C, Allen SW, Trombetta BA, McManus A, Moore MR, Liu J, Cabin DE, Kordasiewicz HB, Mathews J, Arnold SE, Vallabh SM, Minikel EV. Regional variability and genotypic and pharmacodynamic effects on PrP concentration in the CNS. JCI Insight. 2022 Mar 22;7(6):e156532. PMCID: PMC8986079

44. Hughes CS, Foehr S, Garfield DA, Furlong EE, Steinmetz LM, Krijgsveld J. Ultrasensitive proteome analysis using paramagnetic bead technology. Mol Syst Biol. 2014 Oct 30;10(10):757. PMCID: PMC4299378

45. Hughes CS, Moggridge S, Müller T, Sorensen PH, Morin GB, Krijgsveld J. Single-pot, solid-phase-enhanced sample preparation for proteomics experiments. Nat Protoc. 2019 Jan;14(1):68–85. PMID: 30464214

46. Erickson BK, Mintseris J, Schweppe DK, Navarrete-Perea J, Erickson AR, Nusinow DP, Paulo JA, Gygi SP. Active Instrument Engagement Combined with a Real-Time Database Search for Improved Performance of Sample Multiplexing Workflows. J Proteome Res. 2019 Mar 1;18(3):1299–1306. PMCID: PMC7081948

47. Eng JK, McCormack AL, Yates JR. An approach to correlate tandem mass spectral data of peptides with amino acid sequences in a protein database. J Am Soc Mass Spectrom. 1994 Nov;5(11):976–989. PMID: 24226387

48. Huttlin EL, Jedrychowski MP, Elias JE, Goswami T, Rad R, Beausoleil SA, Villén J, Haas W, Sowa ME, Gygi SP. A tissue-specific atlas of mouse protein phosphorylation and expression. Cell. 2010 Dec 23;143(7):1174–1189. PMCID: PMC3035969

49. Elias JE, Gygi SP. Target-decoy search strategy for increased confidence in large-scale protein identifications by mass spectrometry. Nat Methods. 2007 Mar;4(3):207–214. PMID: 17327847

50. Poole W, Gibbs DL, Shmulevich I, Bernard B, Knijnenburg TA. Combining dependent P-values with an empirical adaptation of Brown’s method. Bioinformatics. 2016 Sep 1;32(17):i430–i436. PMCID: PMC5013915

51. Love MI, Huber W, Anders S. Moderated estimation of fold change and dispersion for RNA-seq data with DESeq2. Genome Biol. 2014;15(12):550. PMCID: PMC4302049

52. Mortberg MA, Gentile JE, Nadaf NM, Vanderburg C, Simmons S, Dubinsky D, Slamin A, Maldonado S, Petersen CL, Jones N, Kordasiewicz HB, Zhao HT, Vallabh SM, Minikel EV. A single-cell map of antisense oligonucleotide activity in the brain. Nucleic Acids Res. 2023 May 16;gkad371. PMID: 37188501

